# LamPORE: rapid, accurate and highly scalable molecular screening for SARS-CoV-2 infection, based on nanopore sequencing

**DOI:** 10.1101/2020.08.07.20161737

**Authors:** Phillip James, David Stoddart, Eoghan D Harrington, John Beaulaurier, Lynn Ly, Stuart W Reid, Daniel J Turner, Sissel Juul

**Author notes:** These authors contributed equally (Phillip James, David Stoddart, Eoghan D Harrington, John Beaulaurier). These authors co-supervised the work (Daniel J Turner, Sissel Juul).

## Abstract

LamPORE™ is a rapid way of testing/screening large numbers of samples for the presence or absence of SARS-CoV-2, the virus causing COVID-19. It combines barcoded multi-target amplification, 15-minute barcoded library preparation and real-time nanopore sequencing. Starting with extracted RNA, results can be obtained from 12 samples in approximately an hour and from 96 samples in under 2 hours. High scalability is achieved by combinatorial barcoding.

## Introduction

SARS-CoV-2 emerged in late 2019 and spread rapidly around the world, causing hundreds of thousands of COVID-19-related deaths. The release of the first SARS-CoV-2 genome sequence, on 10^th^ January 2020 (1), allowed the development of tests for the presence or absence of viral RNA from biological samples, which provide a way to identify people who are currently infected by the virus. Although there is some uncertainty about how infectious asymptomatic people are, it is more certain that many people can transmit the virus while being pre-symptomatic, or having mild symptoms (2). As a consequence, rather than only testing people who show symptoms, it is becoming necessary to enable frequent and routine screening of large numbers of people who are not presently showing symptoms, to help return to pre-pandemic activities more safely. For wide-scale screening to be worthwhile, it is important to have assays that are high throughput, accurate and very fast. Epidemiological models show that testing frequency and time-to-result are important components of a surveillance system (3). There is little benefit in being able to screen a large number of samples if the results are not made available quickly enough to inform quarantine decisions or contact tracing. In the US, many labs are taking 5-7 days or more to turn tests around (4).

LamPORE addresses this by combining multiplexed barcoded loop mediated amplification (RT-LAMP), rapid barcoded library preparation and real-time nanopore sequencing to create a way of rapidly testing and screening large numbers of samples for the presence or absence of SARS-CoV-2 RNA. The method fits both large-scale and small-scale laboratory environments.

LAMP is a method of targeted isothermal amplification (5) which can generate micrograms of product from tens of copies of the target region, within 30 minutes at 65°C. Successful amplification is often inferred from a proxy measurement, such as increased turbidity, a colour change or changes in fluorescence. However, although the LAMP reaction itself is very robust, these proxy measurements are less robust and can be affected by substances present in biological samples. It is also not uncommon to see a colour change or increase in turbidity in no-template controls, arising from amplification of primer artefacts, which would lead to a false positive call. Instead of relying on proxy measurements, sequencing can be used as a readout (6). On-target amplification events contain sequences that are not present in the primers and can be identified without ambiguity by alignment. In addition, sequencing provides an opportunity to amplify and detect multiple targets in a single tube.

During nanopore sequencing, an electrical current is measured as template strands pass through each pore on the flow cell array. Conversion of this current into basecalls can start while a strand is translocating. In addition, there is no fixed run time with nanopore sequencing, meaning that the sequencing run can be matched to the data requirements. As a consequence, data analysis can be performed in real time, and results can be returned very rapidly. To maintain these speed advantages, it is necessary to use a correspondingly rapid method of library preparation to convert the amplified products into a form that is compatible with sequencing. Oxford Nanopore Technologies’ (ONT) barcoded rapid library prep kit uses a transposase to convert DNA to a barcoded library that is ready to sequence in approximately 15 minutes, and 96 barcodes are available, allowing prepared samples to be pooled for sequencing. However, by itself this level of multiplexing would be insufficient to satisfy the throughput requirements of frequent testing. It has been shown that a short barcode can be added to one of the primers used in the LAMP reaction (6, 7), and during the course of the reaction the barcode becomes copied multiple times into the products. This raises the possibility of performing dual/combinatorial barcoding without adding additional library preparation steps, thus creating a rapid method of targeted sequencing that is capable of high levels of multiplexing.

In this manuscript, we show that performing a multiplexed amplification reaction, in which three separate regions of the SARS-CoV-2 genome are targeted, performs with high sensitivity (to around Ct37 as measured by RT-qPCR). In addition, the inclusion of a fourth primer set, targeting human actin mRNA allows true negatives to be distinguished from invalid results, where the initial sample was not taken or processed adequately. Suboptimal sampling is suspected to be responsible for false negative results in many SARS-CoV-2 tests (8, 9). Starting with RNA extracted from swabs, results can be obtained from a small number of samples in approximately an hour, and from 96 samples in under 2 hours. This assay is simple to scale from a small number of samples to thousands, with greater degrees of multiplexing achievable by increasing the numbers of LAMP barcodes and / or ONT rapid barcodes.

## Methods

### 1. Amplification and library preparation

Primer sequences for the amplification of three SARS-CoV-2 targets and human actin mRNA were obtained from New England Biolabs (Ipswich, MA, and (10)), and short barcodes were added to the forward inner primers (FIP) as described (6, 7). Primers were synthesised and HPLC-purified by IDT (Coralville, IA). The concentration of actin primers was intentionally lower than for the SARS-CoV-2 primers to prevent amplification of the human target overwhelming any SARS-CoV-2 amplification.

For each FIP barcode, a 10x primer pool was prepared containing each oligonucleotide at the appropriate concentration. Reactions were performed in 96-well plates in such a way that each well in a row received the same barcoded FIP oligo mix, with different barcoded FIPs being used in the different rows. Each LAMP reaction consisted of 25 μl 2x LAMP Master Mix (NEB E1700), 5 μl 10x primer pool and 20 μl RNA sample (or no-template control). Reactions were incubated at 65°C for 35 minutes, followed by 80°C for 5 minutes. Following amplification, reactions were pooled by column, giving 12 pools, each consisting of 8 separate reactions (Fig. 1).

**Figure 1.**
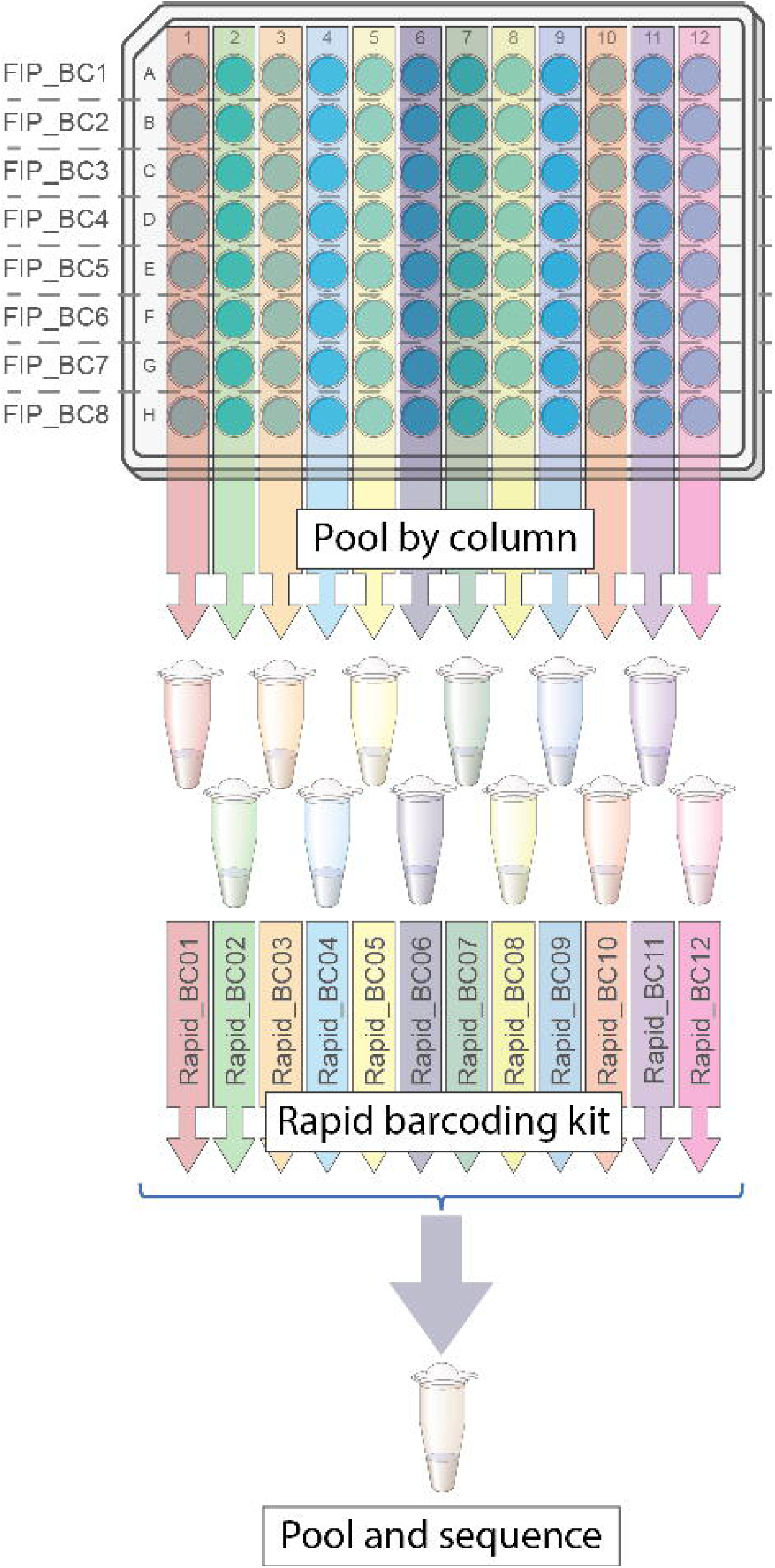
Overview of LamPORE laboratory workflow.

Library preparation was performed separately on each of the 12 pools, in a volume of 10 μl per reaction. Each reaction consisted of 6.5 μl nuclease-free water, 1 μl of pooled LAMP product and 2.5 μl of the appropriate rapid barcode (Oxford Nanopore Technologies, SQK-RBK004). Reactions were mixed and spun down, before being incubated at 30°C for 2 minutes and then 80°C for 2 minutes. All reactions were then pooled into a single 1.5 ml Eppendorf LoBind tube.

The pooled products were purified using 0.8x AMPure beads, were washed in fresh 80% ethanol and were eluted in 15 μl EB buffer. 11 μl of eluate was transferred to a clean 1.5 ml Eppendorf LoBind tube, along with 1 μl rapid adapter (RAP). Reactions were incubated for 5 minutes at room temperature, before being sequenced on a single MinION flow cell for 1 hour, following the manufacturer’s instructions.

### 2. Data analysis

#### i) Barcode and LAMP product identification

In order to call the presence or absence of virus in the sample, the number of reads from each LAMP target must be counted for each sample in the sequencing run. This requires the accurate identification of i) the barcode added during library preparation by the Rapid Barcoding Kit (RBK), ii) the barcode added as part of the FIP oligo during the LAMP reaction and iii) the sequence of the LAMP product associated with each target region.

The RBK barcodes are identified using the guppy_barcoder software (version 4.0.11; command line options “--barcode_kits SQK-RBK004 --detect_mid_strand_barcodes --min_score_mid_barcodes 40”).

The FIP barcode is detected in a two-step process. Firstly, candidate regions are identified by aligning a sequence consisting of the FIP primer with Ns in place of the barcode sequence against all reads using the VSEARCH tool (11) (version 2.14.2; command line options: “--maxaccepts 0 --maxrejects 0 --id 0.75 --strand both --wordlength 5 --minwordmatches 2”). This returns a maximum of 2 candidate regions for each read which are subsequently filtered to remove alignments shorter than 30 nt.

The second step identifies the actual barcode sequence within the candidate region. We selected a strategy to maximise discrimination for these relatively short sequences. Aligning and scoring over the whole candidate region reduced discrimination due to the possibility of sequencing errors in the flanking primer regions. Aligning, or restricting scoring, to only the barcode sequence reduced discrimination due to alignment artefacts around the ends of the barcodes. To avoid such alignment artefacts, whilst maintaining discrimination, 1 nt of the flanking primer sequence was added to each barcode before alignment within the candidate region. Each of the expanded FIP barcode sequences is aligned against the candidate region using the edlib package allowing a maximum edit distance of 1 (12) (version 1.3.8.post1).

The LAMP product associated with each read is identified using the same VSEARCH parameters to align the genome/transcript sequence encompassed spanning the F2-B1 primer locations against each read. A valid LAMP product is detected if the alignment length is greater than 80 nucleotides and greater than 80% identity.

The multimeric nature of the LamPORE reads allows an additional layer of quality control. Each read should only contain sequence from a single LAMP target for a single sample, therefore reads with multiple rapid barcodes, conflicting FIP barcodes or incompatible FIP-product pairings are removed from further consideration. The specific nature of the sequencing analysis allows non-specific amplification, for example primer artefact, to be measured and excluded. Reads with RBK and FIP classifications, but which fail product classification or contain conflicting product regions, are counted as “unclassified”.

#### ii) Determining presence/absence of SARS-CoV-2

Per-sample results of the assay are returned as either positive, negative, inconclusive, or invalid. The calls are made based on the aggregated read counts for each sample across the various targets (i.e. human actin and the three SARS-CoV-2 target regions) and cutoffs were chosen based on 1 hour of sequencing. An invalid call is returned if <50 total classified reads are obtained from across all targets (including both human actin and SARS-CoV-2). A negative call is returned if a sum of <20 reads are obtained from the three SARS-CoV-2 targets (and >= 50 reads in total). An inconclusive call is returned if a sum of >=20 and <50 reads are obtained from the three SARS-CoV-2 targets. A positive call is returned if a sum of >=50 reads are obtained from the three SARS-CoV-2 targets.

#### iii) ROC and F1 score curves

To evaluate the sensitivity and specificity of the assay against the known status of 80 COVID-19 positive clinical RNA samples and a similar number of human RNA-only negatives, we generated receiver operating characteristic (ROC) curves using the metrics.roc_curve function from the scikit-learn package (13). The sum of read counts across each of the three SARS-CoV-2 targets (AS1, E1, and N2) serves as the scoring metric for calling the results positive, negative, inconclusive, or invalid. The ROC curve therefore reveals the sensitivity and specificity of the assay at various thresholding values of that scoring metric. In addition to the curve generated for the SARS-CoV-2 read count sum, we also generated curves for read counts from each individual SARS-CoV-2 target.

The F1 score represents the harmonic mean of the sensitivity and specificity of the assay, defined as 2 * [(1-FPR) * TPR] / [(1-FPR) + TPR], where TPR is the true positive rate and FPR is the false positive rate. We chose the read count threshold (>= 50 total SARS-CoV-2 target reads) in order to maximize the F1 score.

## Results

### i) Assay design

An assay that targeted a single locus from the SARS-CoV-2 genome would potentially lack robustness to sequence variants that occur as the virus evolves. To overcome this, we chose to target three different regions in the SARS-CoV-2 genome, in a single multiplexed reaction. These are ORF1a and the envelope (E) and nucleocapsid (N) genes, with primer sets AS1 (10), E1 and N2 (14), respectively. In addition, as a control for the quality of the initial sample preparation, RNA extraction, reverse transcription and LAMP amplification, we included a set of primers to amplify the human actin mRNA (14). The primers target either side of a splice junction and do not amplify from genomic DNA. As long as the sample has been taken and prepared correctly, actin RNA should be present in all the swab samples, regardless of their SARS-CoV-2 status, and so this provides a way to differentiate between true negatives and invalid samples.

To assess the inclusivity of the triplex SARS-CoV-2 assay, we aligned all primer sequences to the 46,872 human SARS-CoV-2 genomes deposited at GISAID on June 16, 2020 (15, 16). Since not all genomes are high coverage or complete, we excluded 2,105 sequences belonging to 1,939 samples from analysis of at least one primer set because they covered fewer than 90% of all bases in that region. Of the 44,933 genomes with sufficient coverage in all three regions, 2,554 (5.68%) genomes and 179 (0.40%) genomes had a mismatch in one or two primer sets respectively, but a full match for the others. Only 2 (0.004451%) genomes had a mismatch in all three primer sets. The primer sets that we use have a 100% match with most sequences: 97.1% for AS1, 98.7% for E1, and 97.6% for N2. Given the widespread mutations that have been identified in SARS-CoV-2, each primer set has one mismatch for 1.3-2.9% of the strains deposited in GISAID (Table 1). The presence of a single mismatch, however, is unlikely to have a significant impact on the limit of detection, as previously shown in work on MERS-CoV LAMP assays (17).

**Table 1:** *In silico* inclusivity assay.

|  | AS1 | E1 | N2 |
| --- | --- | --- | --- |
| Total Primer Length (nt) | 191 | 168 | 169 |
| Total # of samples evaluated | 45,712 | 46,588 | 46,211 |
| 0 nt mismatches | 44,364 (97.1%) | 45,973 (98.7%) | 45,105 (97.6%) |
| 1 nt mismatch | 1,311 | 602 | 1,050 |
| 2 nt mismatches | 32 | 13 | 48 |
| 3 nt mismatches | 3 | 0 | 4 |
| 4+ nt mismatches | 2 | 0 | 4 |

In order to assess the potential for cross-reactivity with other viruses, we aligned the LAMP primer sequences against sequences of common viruses as well as coronaviruses related to SARS-CoV-2. We determined sequence identity by dividing the sum of aligned primer bases by the sum of primer lengths (Table 2).

**Table 2.** Organisms assessed *in silico* for potential cross-reactivity to the SARS-CoV-2 LamPORE Assay

| Pathogen | GenBank | AS1 (%) | E1 (%) | N2 (%) |
| --- | --- | --- | --- | --- |
| Adenovirus A | NC_001460.1 | 45 | 47.6 | 46.2 |
| Adenovirus B1 | NC_011203.1 | 45.5 | 47 | 49.1 |
| Adenovirus B2 | NC_011202.1 | 44 | 44.6 | 47.9 |
| Adenovirus C | NC_001405.1 | 45 | 45.8 | 45.6 |
| Adenovirus D | NC_010956.1 | 39.8 | 47 | 49.1 |
| Adenovirus E | NC_003266.2 | 39.3 | 45.8 | 45 |
| Adenovirus F | NC_001454.1 | 41.9 | 50 | 42.6 |
| Bordetella pertussis (BPP-1) | NC_005357.1 | 38.2 | 42.9 | 46.7 |
| Candida albicans (L757) | NC_018046.1 | 43.5 | 44.6 | 43.2 |
| Chlamydia pneumoniae | NC_005043.1 | 55.5 | 58.9 | 62.7 |
| Coronavirus 229E | NC_002645.1 | 44 | 50.6 | 49.7 |
| Coronavirus HKU1 | NC_006577.2 | 43.5 | 45.8 | 46.2 |
| Coronavirus NL63 | NC_005831.2 | 45 | 50.6 | 47.3 |
| Coronavirus OC43 | NC_006213.1 | 44.5 | 47.6 | 47.3 |
| Enterovirus D68 | KP745766.1 | 39.8 | 41.7 | 44.4 |
| Haemophilus influenzae | NC_017451.1 | 60.2 | 62.5 | 65.1 |
| Human Metapneumovirus | NC_039199.1 | 40.3 | 45.2 | 43.8 |
| Influenza A (H1N1) | FJ966079.1 | 32.5 | 36.7 | 37.3 |
| Influenza A (H3N2) | KT002533.1 | 34 | 36.9 | 42.6 |
| Influenza B (Victoria) | MN230203.1 | 35.1 | 36 | 36.1 |
| Influenza B (Yamagata) | MK715533.1 | 36.6 | 37.3 | 37.3 |
| Legionella pneumophila | NZ_CP016029.2 | 62.3 | 66.1 | 66.3 |
| MERS-CoV (England 1) | NC_038294.1 | 42.9 | 48.2 | 45 |
| MERS-CoV (HCoV-EMC) | NC_019843.3 | 42.9 | 48.2 | 46.2 |
| Mycoplasma pneumoniae (C267) | NZ_CP014267.1 | 59.7 | 62.5 | 59.2 |
| Pneumocystis jirovecii | NC_020331.1 | 45.5 | 47.6 | 45.6 |
| Pseudomonas aeruginosa | NZ_CP022001.1 | 55.5 | 63.1 | 66.3 |
| Respiratory syncytial virus | NC_001803.1 | 45 | 45.2 | 46.7 |
| Rhinovirus 1 | NC_038311.1 | 38.7 | 38.7 | 44.4 |
| Rhinovirus 14 | NC_001490.1 | 39.3 | 40.5 | 38.5 |
| Rhinovirus C | NC_009996.1 | 36.6 | 41.7 | 43.8 |
| SARS-CoV-1 | NC_004718.3 | 44.5 | **96.4** | 74 |
| SARS-CoV-2 (WU) | MN908947.3 | **100** | **100** | **100** |
| Staphylococcus epidermidis (ATCC 12228) | NZ_CP022247.1 | 65.4 | 65.5 | 60.9 |
| Streptococcus pneumoniae (AP200) | NC_014494.1 | 61.3 | 64.9 | 65.1 |
| Streptococcus pyogenes (AP1) | NZ_CP007537.1 | 62.8 | 63.7 | 60.4 |
| Streptococcus salivarius (YMC-2011) | NC_018285.1 | 43.5 | 51.2 | 47.3 |
| Human parainfluenza 1 | NC_003461.1 | 42.9 | 48.8 | 46.7 |
| Human parainfluenza 2 | NC_003443.1 | 43.5 | 41.7 | 43.8 |
| Human parainfluenza 3 | NC_001796.1 | 42.4 | 46.4 | 45 |
| Human parainfluenza 4 | NC_021928.1 | 41.9 | 48.2 | 45 |
| Tuberculosis (H37Rv) | NC_000962.3 | 53.4 | 58.9 | 61.5 |

SARS-CoV, which is closely related to SARS-CoV-2, was the sole virus to have a match against the total sequence length of the SARS-CoV-2 primers greater than the recommended threshold of 80%. The E-gene primer set has a match >90% with SARS-CoV, but the AS1 and N2 primer sets differ significantly, matching at only 44.5% and 74%, respectively. The likelihood of a false positive is low since SARS-CoV is not known to be in active circulation at present (18). Furthermore, should this situation change, the presence/absence stage of the analysis can be easily modified to identify positive results that are dependent entirely on amplification of the E-gene primer.

### ii) Barcode demultiplexing

LAMP products contain multiple copies of each ∼150 bp target region joined end-to-end, forming strands of up to approximately 5 kb, with consecutive copies of the target region in alternating orientation (Fig. 2a). Following library preparation with the ONT RBK, fragments are reduced to a modal length of around 500 bp, so still typically contain several copies of the target region.

**Figure 2.**
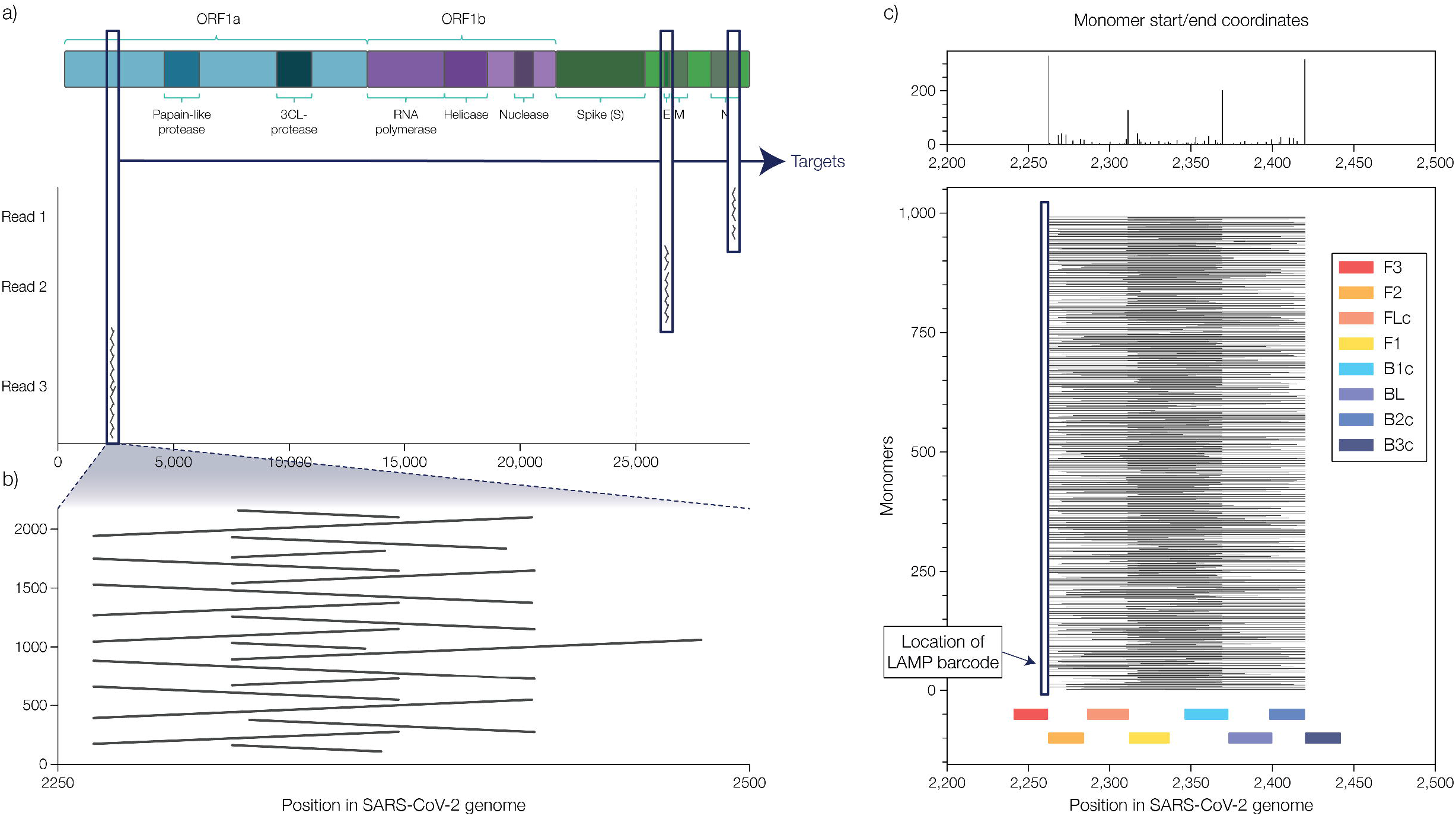
Multimeric reads aligned to the SARS-CoV-2 genome a) reads corresponding to all three assayed loci (genome architecture adapted from (23)) b) a single read aligned to the AS1 target, showing the alternating orientation of unequal consecutive repeating units c) position of LAMP barcode in the repeating units.

More than one forward and reverse primer is used in each LAMP reaction at each target region, so the repeating units are not of a uniform length (Fig. 2b), and because of the location of the barcodes within the FIP oligo, not all copies of the repeating unit contain the LAMP barcodes. This makes it necessary to select reads that do contain LAMP barcodes (Fig. 2c). All LamPORE reads contain an ONT barcode at one of the ends, and by selecting for LAMP barcodes, we typically retain the approximately 70% of reads which thus contain both barcodes and the target region.

Barcode specificity was assessed by inspection of counts in the negative control wells of the clinical sample plates (described below). After excluding known contamination, the average number of counts attributed to SARS-Cov-2 targets in negative control wells was 1.6 +/- 0.2 (standard error on mean). The highest number of counts was 9. These counts were well below the “inconclusive” call threshold

### iii) Primer artefacts

Primer artefacts can accumulate during the LAMP reaction, and as a result, the consequence of judging successful amplification by a proxy measurement, such as a colour change or increase in turbidity, can be a false positive call. This is avoided when sequencing is used as a readout: reads are aligned to a reference sequence, and for a read to be considered valid, it must consist of inverted repeats of large stretches of the target region, including target-specific sequences present that do not exist in the primers. Alignments of valid reads are contiguous across the majority of the target region (Fig. 3a). In contrast, primer artefacts consist entirely of sequences covered by the primers, and these tend to align as short segments interspersed with gaps.

**Figure 3.**
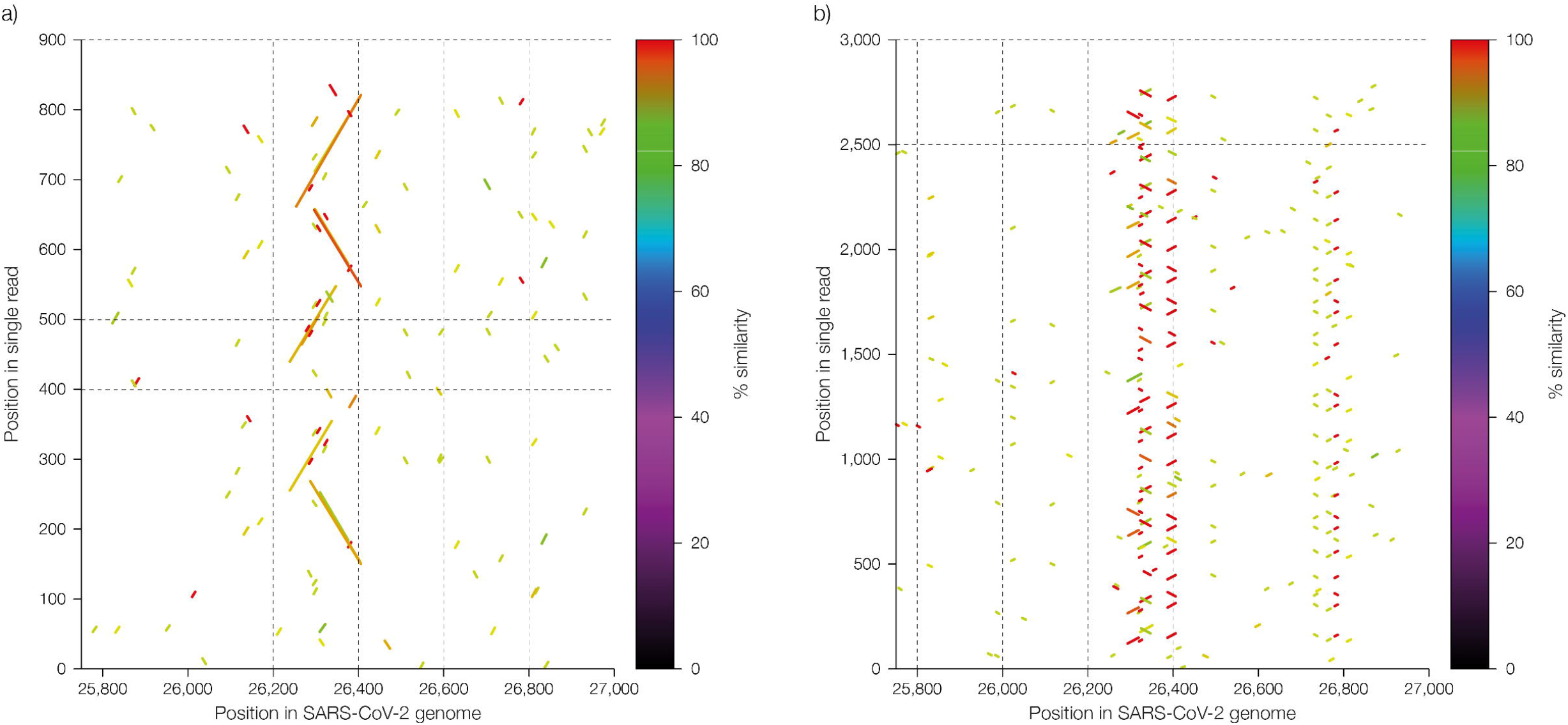
Distinguishing between valid reads and primer artefacts by alignment a) valid reads consist of inverted repeats that align across the majority of the target region, whereas b) primer artefacts align as short segments interspersed with gaps.

### iv) FIP barcode optimisation

Verification of the FIP barcodes for each target was carried out using a dilution series of the Twist Synthetic RNA Control 2 (Twist Biosciences) for the SARS-CoV-2 loci and total human RNA extracted from GM12878 (Coriell) for the actin control. Template quantities ranged from 20-250 copies per reaction. We observed that not only does the presence of the barcode influence the sensitivity of the reaction, the sequence of the barcode also affects performance, with some barcoded FIPs working with higher sensitivity than others. We excluded the worst-performing barcoded FIPs and in this way we reduced our initial 12 barcodes to the best-performing 8, all of which were capable of amplifying from 20 copies in a 50 μl LAMP reaction (Fig. 4). When used in combination with 12 rapid barcodes, 96 combinations are produced.

**Figure 4.**
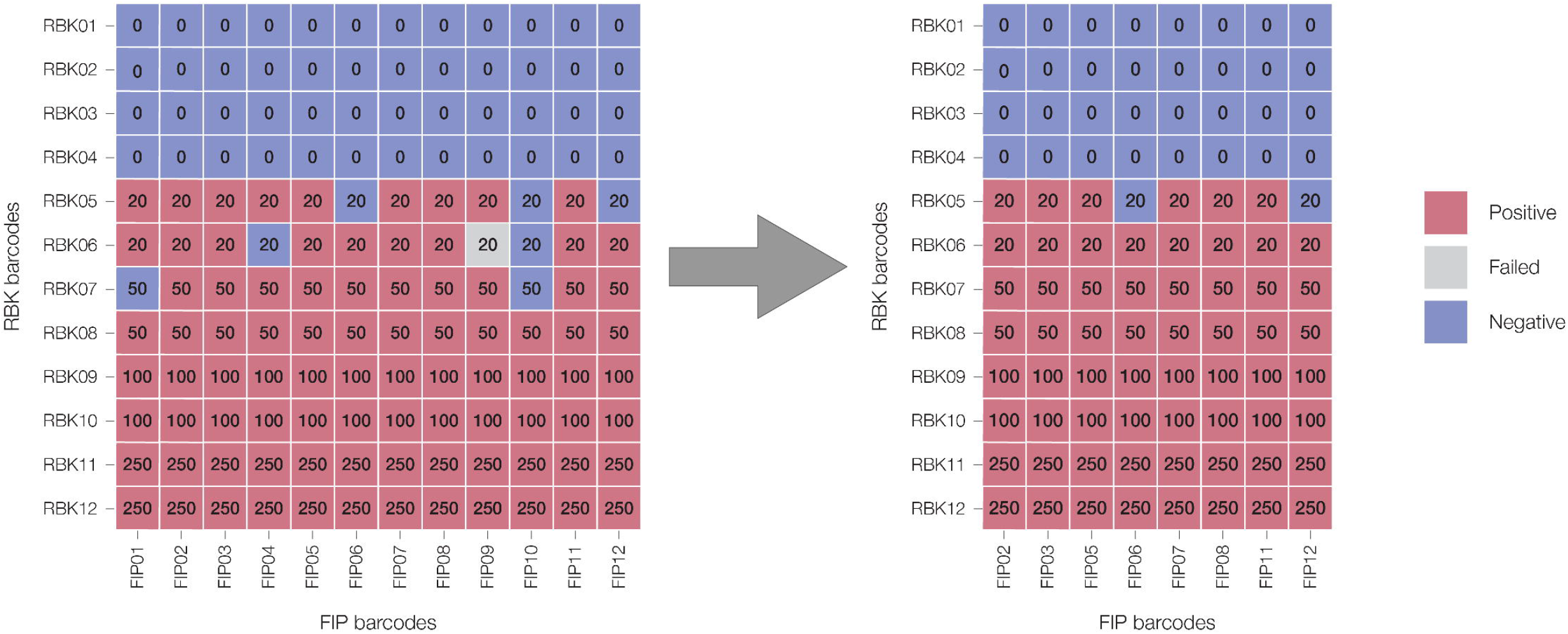
Selection of the best performing FIP barcoded primers from a larger number of candidate oligos. Numbers indicate the quantity of template copies added to each reaction.

### v) Clinical samples

To expand our evaluation of the assay’s performance, we obtained 80 clinical samples, consisting of RNA which had been extracted from nasopharyngeal swabs. The samples had been found to be positive for SARS-CoV-2 RNA by RT-qPCR (19), and spanned a range of Ct values, from Ct = 19 for the highest viral load to Ct = 38 for the lowest. In the absence of RT-qPCR-verified negative samples, we prepared a similar number of reaction negatives using total human RNA. A sufficient number of sequences corresponding to the actin control fragment were obtained in all negative samples for these to be called as valid, and in 81 out of 85 samples, a negative call was obtained. Read-count results indicate the amplification of targets E1 or N2 in the four positives was due to contamination. Out of the 80 RT-qPCR-verified positives, 79 were called positive in the LamPORE analysis. The false negative corresponded to the lowest Ct sample, Ct = 38. The two samples at Ct = 37 were called positive (Table 3 and Supplementary Table 1).

**Table 3.** Representative selection of results obtained from performing LamPORE on clinical extracts, which had been validated as positive by RT-qPCR. For full results, see Supplementary Table 1.

| Sample ID | RT-qPCR Ct | Actin | AS1 | E1 | N2 | Unclassified | Call | True status |
| --- | --- | --- | --- | --- | --- | --- | --- | --- |
| ONT5555 | 18 | 1 | 725 | 208 | 110 | 187 | POS | Positive |
| ONT1427 | 20 | 0 | 1025 | 37 | 33 | 169 | POS | Positive |
| ONT6807 | 22 | 1 | 1511 | 259 | 252 | 296 | POS | Positive |
| ONT9138 | 22 | 0 | 1634 | 302 | 285 | 325 | POS | Positive |
| ONT3768 | 22 | 2 | 1865 | 164 | 191 | 378 | POS | Positive |
| ONT9941 | 25 | 0 | 291 | 17 | 13 | 51 | POS | Positive |
| ONT2659 | 25 | 5 | 3504 | 91 | 74 | 1115 | POS | Positive |
| ONT6574 | 25 | 2 | 1229 | 61 | 54 | 306 | POS | Positive |
| ONT9410 | 26 | 1 | 1333 | 155 | 93 | 323 | POS | Positive |
| ONT0371 | 26 | 2 | 1016 | 20 | 20 | 193 | POS | Positive |
| ONT0844 | 28 | 0 | 1028 | 20 | 23 | 180 | POS | Positive |
| ONT7273 | 29 | 0 | 1257 | 1 | 3 | 163 | POS | Positive |
| ONT9661 | 29 | 1 | 550 | 0 | 7 | 92 | POS | Positive |
| ONT7343 | 30 | 3 | 1199 | 14 | 25 | 236 | POS | Positive |
| ONT2196 | 31 | 2 | 173 | 1 | 0 | 29 | POS | Positive |
| ONT7466 | 32 | 1 | 1369 | 1 | 2 | 202 | POS | Positive |
| ONT3588 | 32 | 13 | 2155 | 45 | 10 | 308 | POS | Positive |
| ONT6853 | 33 | 1 | 257 | 0 | 0 | 37 | POS | Positive |
| ONT7433 | 36 | 0 | 608 | 1 | 2 | 151 | POS | Positive |
| ONT1196 | 36 | 7 | 222 | 0 | 1 | 38 | POS | Positive |
| Human RNA | N/A | 538 | 7 | 0 | 0 | 131 | NEG | Negative |
| Human RNA | N/A | 1209 | 3 | 0 | 0 | 254 | NEG | Negative |
| Human RNA | N/A | 626 | 1 | 0 | 0 | 137 | NEG | Negative |
| Human RNA | N/A | 58 | 0 | 0 | 1 | 18 | NEG | Negative |
| Human RNA | N/A | 211 | 4 | 0 | 0 | 49 | NEG | Negative |

### vi) Assay sensitivity and specificity

ROC curves generated from 80 COVID-19 positive clinical samples and 85 COVID-19 human RNA negatives show good concordance between SARS-CoV-2 detection via the LamPORE assay and the RT-qPCR verified status, with an area under the curve (AUC) of 0.993 for the metric used in calling the results (sum of SARS-CoV-2 target reads, Fig. 5a). We correctly called 79 samples positive across the 80 COVID-19 positive samples at our optimal read count threshold. Read count results suggest that contamination led to the amplification of the SARS-CoV-2 targets E1 or N2 in the four samples that generated false positive calls. The optimal read count threshold of >=50 reads for a positive call was selected by maximizing the F1 score corresponding to the ROC curves (Fig. 5b).

**Figure 5.**
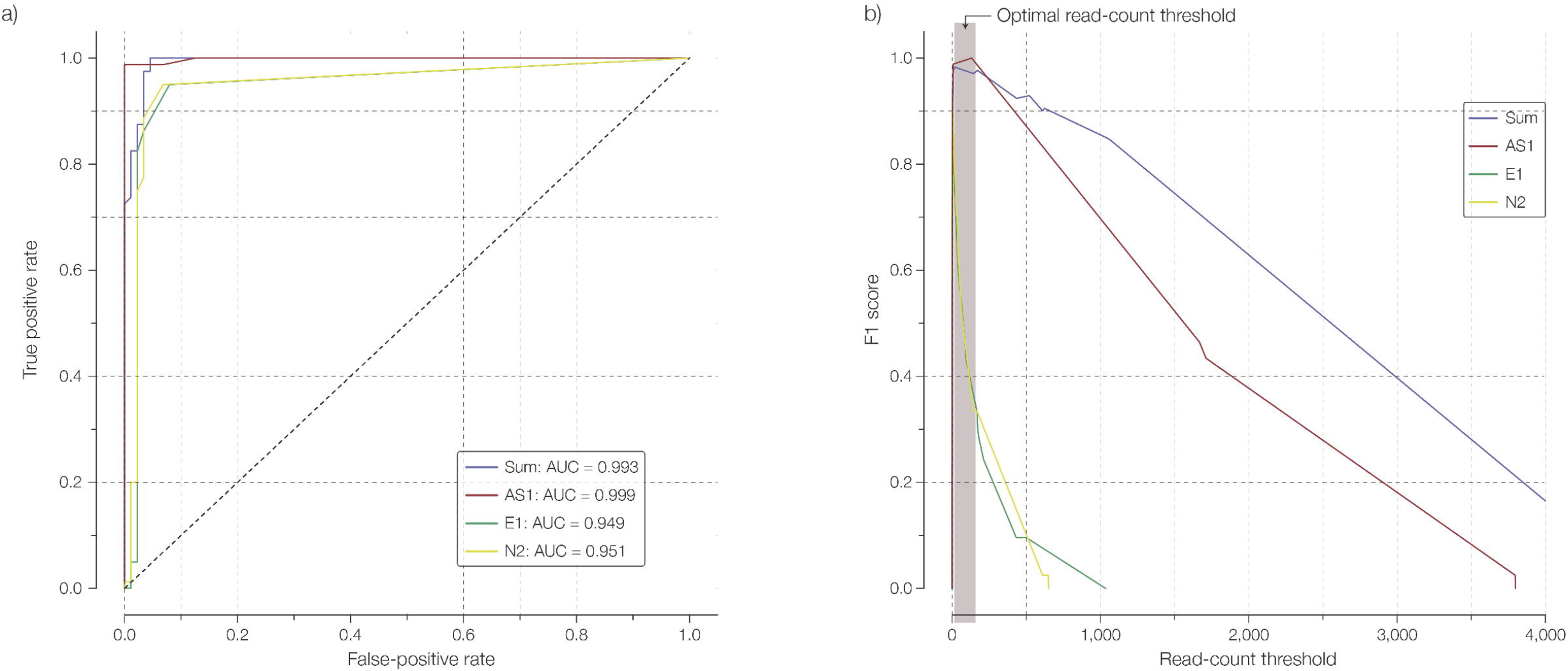
Assay performance and threshold selection. (a) The ROC curve showing the true and false positive rates at varying SARS-CoV-2 target read count thresholds, for the sum of read counts from all three SARS-CoV-2 targets and each individual SARS-CoV-2 target. (b) The F1 score was therefore used to identify the optimal read count threshold for calling a SARS-CoV-2 positive sample.

## Discussion

In this manuscript we present a method which combines the rapid target-specific amplification provided by LAMP, a method of transposase-based library preparation, and real-time nanopore sequencing and data analysis. The resulting combination, LamPORE, is rapid, sensitive and highly scalable and here we demonstrate LamPORE’s efficacy for detecting the presence or absence of SARS-CoV-2 RNA in clinical samples. Studies using much larger sample sets have been recently conducted to establish diagnostic performance claims (20).

The end-to-end procedure, beginning with 96 RNA extracts, and ending with positive and negative calls, can be performed in under 2 hours when sequencing on a MinION™ or GridION™. The number of samples that can be sequenced in parallel can be increased by expanding either the number of LAMP barcodes or the number of ONT rapid barcodes. In these circumstances, it is necessary to extend the length of the sequencing run. When using 12 different LAMP barcodes combined with 96 rapid barcodes (= 1,152 samples) we found that 4 hours of MinION sequencing was sufficient. After sequencing it is possible to remove the sample strands from the flow cell with a nuclease flush, and to load a fresh set of samples. Having a larger number of pores per flow cell, the length of the corresponding sequencing run is shorter on the PromethION, but alternatively, it is possible to use larger multiplexes.

The remaining bottleneck in the current end-to-end workflow is that of extracting RNA from the biological samples. Recent publications have indicated that saliva is a suitable source of SARS-CoV-2 RNA in infected patients (21, 22), and we have found that following heat-treatment, saliva with spiked-in inactivated SARS-Cov-2 virions can be amplified and sequenced successfully. This is currently being investigated further.

We have seen in the work presented here that LAMP is capable of amplifying several targets simultaneously. LamPORE relies on sequencing, as opposed to a colour change, which raises the possibility of using a single multiplexed LamPORE reaction to detect many different pathogens. In the case of co-infection, it should also be possible to identify which combination of pathogens is present.

## Data Availability

The nucleotide sequences of the SARS-CoV-2 genomes used in the primer inclusivity and cross-reactivity analysis are available, upon free registration, from the GISAID database.

https://www.gisaid.org/

## Data availability

The nucleotide sequences of the SARS-CoV-2 genomes used in the primer inclusivity and cross-reactivity analysis are available, upon free registration, from the GISAID database (https://www.gisaid.org/).

## Acknowledgements

We would like to thank Dr. Thushan de Silva, University of Sheffield, UK and Dr. Cariad Evans, Sheffield Teaching Hospitals NHS Foundation Trust, UK for providing the positive, RT-qPCR-verified SARS-CoV-2 samples. We are also grateful to Adam Peltan and Nathan Tanner at New England Biolabs for discussions about LAMP assays and their help with primer sequences.

We gratefully acknowledge the authors from the originating laboratories responsible for obtaining the specimens, and the submitting laboratories where genetic sequence data were generated and shared via the GISAID Initiative, on which some of this research is based.

## Regulatory status

Oxford Nanopore Diagnostics have CE marked LamPORE for diagnostic use. Other regulatory submissions are being pursued

## Ethics declaration

Patient consent was not required as these were routinely collected samples and stored as per normal protocols used for clinical practice. They did not contain any cellular material and were therefore not subject to HTA. Therefore, as for assay development, using anonymised samples, no IRAS/ethics was required or sought.

