## Supplementary Table 1 for "LamPORE: rapid, accurate and highly scalable molecular screening for SARS-CoV-2 infection, based on nanopore sequencing"

| Sample ID | RT-qPCR Ct | Actin | AS1 | E1 | N2 | Unclassified | Call | True status |
|---|---|---|---|---|---|---|---|---|
| Human RNA | N/A | 58 | 0 | 0 | 1 | 18 | NEG | Negative |
| Human RNA | N/A | 59 | 0 | 0 | 0 | 12 | NEG | Negative |
| Human RNA | N/A | 59 | 0 | 0 | 0 | 19 | NEG | Negative |
| Human RNA | N/A | 65 | 1 | 0 | 0 | 27 | NEG | Negative |
| Human RNA | N/A | 65 | 1 | 0 | 0 | 17 | NEG | Negative |
| Human RNA | N/A | 73 | 0 | 0 | 0 | 15 | NEG | Negative |
| Human RNA | N/A | 79 | 0 | 0 | 0 | 28 | NEG | Negative |
| Human RNA | N/A | 99 | 0 | 0 | 0 | 34 | NEG | Negative |
| Human RNA | N/A | 109 | 0 | 0 | 0 | 21 | NEG | Negative |
| Human RNA | N/A | 115 | 2 | 0 | 0 | 47 | NEG | Negative |
| Human RNA | N/A | 119 | 1 | 0 | 0 | 43 | NEG | Negative |
| Human RNA | N/A | 120 | 1 | 0 | 0 | 30 | NEG | Negative |
| Human RNA | N/A | 138 | 1 | 0 | 0 | 27 | NEG | Negative |
| Human RNA | N/A | 161 | 2 | 0 | 0 | 70 | NEG | Negative |
| Human RNA | N/A | 164 | 0 | 0 | 0 | 79 | NEG | Negative |
| Human RNA | N/A | 169 | 1 | 0 | 0 | 44 | NEG | Negative |
| Human RNA | N/A | 181 | 5 | 0 | 0 | 63 | NEG | Negative |
| Human RNA | N/A | 185 | 1 | 1 | 7 | 456 | NEG | Negative |
| Human RNA | N/A | 193 | 1 | 0 | 0 | 55 | NEG | Negative |
| Human RNA | N/A | 199 | 0 | 0 | 0 | 62 | NEG | Negative |
| Human RNA | N/A | 206 | 0 | 0 | 0 | 71 | NEG | Negative |
| Human RNA | N/A | 211 | 4 | 0 | 0 | 49 | NEG | Negative |
| Human RNA | N/A | 212 | 0 | 0 | 0 | 66 | NEG | Negative |
| Human RNA | N/A | 214 | 1 | 0 | 0 | 45 | NEG | Negative |
| Human RNA | N/A | 221 | 1 | 0 | 597 | 227 | POS | Negative |
| Human RNA | N/A | 232 | 0 | 0 | 0 | 89 | NEG | Negative |
| Human RNA | N/A | 236 | 4 | 0 | 0 | 88 | NEG | Negative |
| Human RNA | N/A | 240 | 0 | 0 | 0 | 55 | NEG | Negative |
| Human RNA | N/A | 243 | 0 | 0 | 0 | 89 | NEG | Negative |
| Human RNA | N/A | 245 | 0 | 0 | 0 | 53 | NEG | Negative |
| Human RNA | N/A | 253 | 0 | 0 | 0 | 98 | NEG | Negative |
| Human RNA | N/A | 261 | 0 | 0 | 0 | 52 | NEG | Negative |
| Human RNA | N/A | 262 | 3 | 0 | 0 | 47 | NEG | Negative |
| Human RNA | N/A | 272 | 5 | 0 | 0 | 54 | NEG | Negative |
| Human RNA | N/A | 273 | 6 | 0 | 0 | 54 | NEG | Negative |
| Human RNA | N/A | 274 | 0 | 0 | 0 | 54 | NEG | Negative |
| Human RNA | N/A | 282 | 1 | 0 | 0 | 65 | NEG | Negative |
| Human RNA | N/A | 306 | 0 | 0 | 0 | 87 | NEG | Negative |
| Human RNA | N/A | 326 | 5 | 0 | 0 | 90 | NEG | Negative |
| Human RNA | N/A | 332 | 0 | 0 | 1 | 80 | NEG | Negative |
| Human RNA | N/A | 336 | 1 | 0 | 0 | 79 | NEG | Negative |
| Human RNA | N/A | 337 | 0 | 0 | 0 | 79 | NEG | Negative |

| | | | | | | | | |
|---|---|---|---|---|---|---|---|---|
| Human RNA | N/A | 369 | 1 | 0 | 0 | 63 | NEG | Negative |
| Human RNA | N/A | 395 | 2 | 2 | 0 | 127 | NEG | Negative |
| Human RNA | N/A | 406 | 0 | 1 | 0 | 98 | NEG | Negative |
| Human RNA | N/A | 438 | 4 | 1 | 0 | 182 | NEG | Negative |
| Human RNA | N/A | 442 | 0 | 0 | 0 | 204 | NEG | Negative |
| Human RNA | N/A | 472 | 1 | 0 | 0 | 239 | NEG | Negative |
| Human RNA | N/A | 479 | 1 | 0 | 0 | 129 | NEG | Negative |
| Human RNA | N/A | 481 | 2 | 424 | 0 | 3027 | POS | Negative |
| Human RNA | N/A | 490 | 4 | 0 | 0 | 113 | NEG | Negative |
| Human RNA | N/A | 490 | 1 | 0 | 139 | 169 | POS | Negative |
| Human RNA | N/A | 506 | 7 | 0 | 0 | 125 | NEG | Negative |
| Human RNA | N/A | 507 | 3 | 1014 | 0 | 389 | POS | Negative |
| Human RNA | N/A | 522 | 1 | 0 | 0 | 334 | NEG | Negative |
| Human RNA | N/A | 523 | 5 | 0 | 0 | 104 | NEG | Negative |
| Human RNA | N/A | 538 | 7 | 0 | 0 | 131 | NEG | Negative |
| Human RNA | N/A | 544 | 5 | 3 | 0 | 165 | NEG | Negative |
| Human RNA | N/A | 549 | 0 | 0 | 0 | 130 | NEG | Negative |
| Human RNA | N/A | 564 | 2 | 0 | 0 | 162 | NEG | Negative |
| Human RNA | N/A | 586 | 0 | 0 | 0 | 160 | NEG | Negative |
| Human RNA | N/A | 600 | 0 | 0 | 0 | 121 | NEG | Negative |
| Human RNA | N/A | 603 | 0 | 0 | 0 | 303 | NEG | Negative |
| Human RNA | N/A | 614 | 2 | 0 | 0 | 137 | NEG | Negative |
| Human RNA | N/A | 626 | 1 | 0 | 0 | 137 | NEG | Negative |
| Human RNA | N/A | 628 | 0 | 1 | 0 | 261 | NEG | Negative |
| Human RNA | N/A | 652 | 2 | 1 | 0 | 157 | NEG | Negative |
| Human RNA | N/A | 702 | 0 | 0 | 0 | 175 | NEG | Negative |
| Human RNA | N/A | 706 | 0 | 0 | 0 | 189 | NEG | Negative |
| Human RNA | N/A | 737 | 1 | 0 | 0 | 135 | NEG | Negative |
| Human RNA | N/A | 751 | 4 | 0 | 0 | 931 | NEG | Negative |
| Human RNA | N/A | 754 | 2 | 0 | 0 | 166 | NEG | Negative |
| Human RNA | N/A | 783 | 0 | 0 | 0 | 182 | NEG | Negative |
| Human RNA | N/A | 1143 | 0 | 0 | 0 | 355 | NEG | Negative |
| Human RNA | N/A | 1157 | 0 | 0 | 0 | 230 | NEG | Negative |
| Human RNA | N/A | 1209 | 3 | 0 | 0 | 254 | NEG | Negative |
| Human RNA | N/A | 1313 | 0 | 0 | 0 | 342 | NEG | Negative |
| Human RNA | N/A | 1359 | 1 | 0 | 0 | 287 | NEG | Negative |
| Human RNA | N/A | 1447 | 0 | 0 | 0 | 346 | NEG | Negative |
| Human RNA | N/A | 1490 | 1 | 0 | 1 | 276 | NEG | Negative |
| Human RNA | N/A | 1538 | 0 | 0 | 0 | 453 | NEG | Negative |
| Human RNA | N/A | 1655 | 2 | 0 | 0 | 361 | NEG | Negative |
| Human RNA | N/A | 1725 | 0 | 0 | 0 | 381 | NEG | Negative |
| Human RNA | N/A | 1832 | 2 | 0 | 0 | 524 | NEG | Negative |
| Human RNA | N/A | 1908 | 3 | 0 | 0 | 493 | NEG | Negative |

| | | | | | | | | |
|---|---|---|---|---|---|---|---|---|
| ONT6807 | 22 | 1 | 1511 | 259 | 252 | 296 | POS | Positive |
| ONT9138 | 22 | 0 | 1634 | 302 | 285 | 325 | POS | Positive |
| ONT1427 | 20 | 0 | 1025 | 37 | 33 | 169 | POS | Positive |
| ONT7343 | 30 | 3 | 1199 | 14 | 25 | 236 | POS | Positive |
| ONT9410 | 26 | 1 | 1333 | 155 | 93 | 323 | POS | Positive |
| ONT0371 | 26 | 2 | 1016 | 20 | 20 | 193 | POS | Positive |
| ONT6853 | 33 | 1 | 257 | 0 | 0 | 37 | POS | Positive |
| ONT7273 | 29 | 0 | 1257 | 1 | 3 | 163 | POS | Positive |
| ONT2196 | 31 | 2 | 173 | 1 | 0 | 29 | POS | Positive |
| ONT7433 | 36 | 0 | 608 | 1 | 2 | 151 | POS | Positive |
| ONT0844 | 28 | 0 | 1028 | 20 | 23 | 180 | POS | Positive |
| ONT9941 | 25 | 0 | 291 | 17 | 13 | 51 | POS | Positive |
| ONT7466 | 32 | 1 | 1369 | 1 | 2 | 202 | POS | Positive |
| ONT1196 | 36 | 7 | 222 | 0 | 1 | 38 | POS | Positive |
| ONT9661 | 29 | 1 | 550 | 0 | 7 | 92 | POS | Positive |
| ONT5555 | 18 | 1 | 725 | 208 | 110 | 187 | POS | Positive |
| ONT3768 | 22 | 2 | 1865 | 164 | 191 | 378 | POS | Positive |
| ONT3588 | 32 | 13 | 2155 | 45 | 10 | 308 | POS | Positive |
| ONT2659 | 25 | 5 | 3504 | 91 | 74 | 1115 | POS | Positive |
| ONT6574 | 25 | 2 | 1229 | 61 | 54 | 306 | POS | Positive |
| ONT9590 | 29 | 7 | 1812 | 49 | 55 | 280 | POS | Positive |
| ONT4401 | 19 | 1 | 367 | 11 | 43 | 72 | POS | Positive |
| ONT3660 | 33 | 1 | 129 | 0 | 0 | 15 | POS | Positive |
| ONT3273 | 30 | 2 | 313 | 7 | 9 | 77 | POS | Positive |
| ONT9911 | 20 | 2 | 3720 | 278 | 352 | 802 | POS | Positive |
| ONT2355 | 28 | 2 | 162 | 2 | 4 | 36 | POS | Positive |
| ONT2843 | 24 | 2 | 290 | 5 | 7 | 100 | POS | Positive |
| ONT4252 | 26 | 0 | 566 | 7 | 13 | 141 | POS | Positive |
| ONT6844 | 27 | 4 | 3475 | 38 | 55 | 517 | POS | Positive |
| ONT7684 | 33 | 4 | 1378 | 1 | 2 | 250 | POS | Positive |
| ONT0939 | 20 | 0 | 1759 | 125 | 116 | 773 | POS | Positive |
| ONT2325 | 23 | 2 | 1724 | 86 | 119 | 798 | POS | Positive |
| ONT8376 | 31 | 1 | 767 | 7 | 4 | 102 | POS | Positive |
| ONT7008 | 37 | 26 | 445 | 14 | 51 | 351 | POS | Positive |
| ONT0821 | 28 | 4 | 1215 | 62 | 125 | 192 | POS | Positive |
| ONT1078 | 35 | 0 | 548 | 6 | 15 | 84 | POS | Positive |
| ONT0745 | 29 | 1 | 897 | 47 | 37 | 149 | POS | Positive |
| ONT1833 | 27 | 0 | 1527 | 7 | 33 | 215 | POS | Positive |
| ONT0293 | 34 | 3 | 1881 | 1 | 4 | 274 | POS | Positive |
| ONT3580 | 29 | 0 | 1922 | 2 | 14 | 279 | POS | Positive |
| ONT7926 | 33 | 40 | 1856 | 24 | 57 | 484 | POS | Positive |
| ONT6045 | 29 | 0 | 1072 | 1 | 1 | 130 | POS | Positive |
| ONT8025 | 20 | 4 | 1351 | 126 | 213 | 263 | POS | Positive |

| | | | | | | | | |
|---|---|---|---|---|---|---|---|---|
| ONT5242 | 22 | 0 | 1202 | 338 | 132 | 260 | POS | Positive |
| ONT8738 | 30 | 47 | 1232 | 38 | 11 | 212 | POS | Positive |
| ONT4776 | 28 | 6 | 1420 | 169 | 87 | 272 | POS | Positive |
| ONT0117 | 38 | 110 | 4 | 12 | 1 | 61 | NEG | Positive |
| ONT3953 | 19 | 1 | 2193 | 531 | 492 | 555 | POS | Positive |
| ONT1173 | 21 | 2 | 3200 | 490 | 636 | 660 | POS | Positive |
| ONT7580 | 25 | 0 | 1242 | 3 | 22 | 176 | POS | Positive |
| ONT3634 | 25 | 3 | 1714 | 258 | 177 | 309 | POS | Positive |
| ONT5038 | 26 | 0 | 975 | 120 | 39 | 178 | POS | Positive |
| ONT5904 | 22 | 0 | 1556 | 27 | 91 | 264 | POS | Positive |
| ONT6871 | 37 | 3 | 1155 | 17 | 22 | 493 | POS | Positive |
| ONT1866 | 32 | 24 | 764 | 27 | 1 | 170 | POS | Positive |
| ONT0343 | 21 | 1 | 1526 | 50 | 40 | 198 | POS | Positive |
| ONT3704 | 30 | 2 | 1074 | 54 | 25 | 163 | POS | Positive |
| ONT5658 | 36 | 40 | 1238 | 86 | 37 | 193 | POS | Positive |
| ONT7602 | 26 | 0 | 1441 | 105 | 89 | 213 | POS | Positive |
| ONT0262 | 27 | 1 | 1292 | 51 | 77 | 224 | POS | Positive |
| ONT4895 | 23 | 0 | 628 | 10 | 27 | 107 | POS | Positive |
| ONT0547 | 15 | 0 | 1265 | 10 | 37 | 178 | POS | Positive |
| ONT9852 | 18 | 0 | 1500 | 538 | 404 | 420 | POS | Positive |
| ONT0081 | 21 | 0 | 615 | 10 | 6 | 94 | POS | Positive |
| ONT8130 | 20 | 2 | 1773 | 10 | 72 | 253 | POS | Positive |
| ONT0697 | 36 | 4 | 1802 | 2 | 0 | 191 | POS | Positive |
| ONT7531 | 22 | 0 | 1465 | 164 | 164 | 300 | POS | Positive |
| ONT0047 | 31 | 1 | 2356 | 31 | 23 | 301 | POS | Positive |
| ONT1538 | 20 | 0 | 1309 | 36 | 176 | 241 | POS | Positive |
| ONT5522 | 27 | 0 | 1510 | 179 | 175 | 316 | POS | Positive |
| ONT8021 | 32 | 1 | 865 | 5 | 1 | 109 | POS | Positive |
| ONT8526 | 28 | 0 | 1004 | 7 | 6 | 147 | POS | Positive |
| ONT8889 | 33 | 9 | 2726 | 55 | 42 | 409 | POS | Positive |
| ONT8609 | 22 | 2 | 1634 | 329 | 326 | 434 | POS | Positive |
| ONT0409 | 23 | 1 | 3500 | 673 | 339 | 584 | POS | Positive |
| ONT7628 | 23 | 2 | 1449 | 179 | 207 | 286 | POS | Positive |
| ONT4880 | 31 | 2 | 1677 | 27 | 36 | 226 | POS | Positive |
| ONT8057 | 26 | 0 | 1726 | 32 | 10 | 201 | POS | Positive |
| ONT1353 | 30 | 0 | 1631 | 14 | 9 | 267 | POS | Positive |
| ONT6461 | 26 | 0 | 2211 | 20 | 203 | 344 | POS | Positive |
